# Response of cell-free miRNAs to acute exercise: A systematic Review and Meta-Analysis

**DOI:** 10.1101/2022.12.20.22283756

**Authors:** Kirstin MacGregor, Colin N Moran, Sophie Broome, Patrick J Owen, Séverine Lamon, Danielle Hiam

## Abstract

**Aim:** Cell-free microRNAs (cf-miRNAs) are secreted from cells and transported via the blood to exert effects on target tissues. Numerous pathophysiological adaptations, including exercise, alter cf-miRNA levels. In this systematic review we examined miRNA expression at multiple timepoints following acute exercise to capture the temporal dynamics of the response.

**Method:** The systematic review was registered in PROSPERO (CRD42021256303). A multi-level meta-analysis was used to compare the changes in cf-miRNA levels and the influence of exercise modality. An exploratory machine-learning-based approach was used to capture influential moderators. Primary studies were retrieved from PubMed, Medline Complete, CINAHL and SPORTDiscuss from inception to 12 August 2025. Relative changes in cf-miRNA expression in response to exercise were computed. The ROBINS-I, GRADE, and AMSTAR2 tools were used to assess risk of bias, strength of evidence and methodological quality of the systematic review respectively.

**Results:** Thirty-five studies that included an acute exercise intervention in healthy (N=736) males and females aged 18–45 years met the eligibility criteria. Muscle enriched cf-miR-1 (n=373), cf-miR-133a (n=373) and cf-miR-133b (n=295) levels increased (logFC [95%CI]) 1–2 hours (cf-miR1: 0.92 [0.14, 1.69]; cf-miR133a: 0.86 [0.45, 1.26]; cf-miR-133b: 0.94 [0.16, 1.72]) and 24 hours post-exercise (cf-miR1: 0.64 [0.03, 1.24]; cf-miR133a: 0.5 [0.16, 0.84]; cf-miR-133b: 0.44 [0.02, 0.86]). Interestingly, there were no differences in the overall expression levels of the included miRNAs between exercise modality groups.

**Conclusion:** Muscle enriched cf-miRNAs are dynamically regulated post-exercise, displaying distinct temporal expression patterns and provide critical insights to guide future research on cf-miRNA dynamics in exercise contexts.

**Key Points:**

- One of the first studies to apply a multi-level meta-analysis in cf-miRNA research, capturing the temporal dynamics of cf-miRNA responses to acute exercise.
- Acute exercise triggers distinct, time-dependent changes in cf-miRNA levels, with muscle-enriched miR-1, -133a, and -133b upregulated, and cf-miR-20a and -486 downregulated post-exercise.
- Findings are limited by small sample sizes and methodological variability across studies, underscoring the need for more rigorous and consistent research designs

## Introduction

Extra-cellular or cell-free microRNAs (cf-miRNAs) are circulating miRNA molecules found in most biological fluids including blood, urine and saliva [1] that mediate protein expression via the regulation of translation [2-4]. Cf-miRNAs are secreted from cells [5] and, while our understanding of their role and relevance in circulation is limited, specific cf-miRNAs display high levels of regulation in numerous pathological [6] and physiological conditions, including exercise [7].

Exercise is a potent physiological stressor triggering acute and chronic biological responses across the cardiovascular, musculoskeletal, and endocrine systems. Exercise relies on the activation of oxygen-dependent or –independent metabolic pathways to trigger muscle contraction [8] and results in a range of positive adaptations at the systemic and cellular levels. Depending on the type of exercise performed, long term adaptations include muscle hypertrophy and improvements in cardiovascular output, muscle fatigability and metabolic function. From a molecular perspective, even a single bout of exercise is enough to elicit a rapid physiological response in the muscle [9, 10] and the heart [11], at the gene [9, 10], protein [9, 12] and miRNA level [12, 13].

Upon acute physiological stress, such as exercise, miRNAs produced in the muscle and other tissues are selectively or non-selectively secreted into the circulation [5]. This might be to exert their effects on recipient cells [14], underpinning a role in cross-tissue communication [14, 15]. Plasma and serum constitute readily available biological miRNA pools that underlie the popularity of cf-miRNA studies and the exponential increase of related research outputs [16]. Research into cf-miRNAs and exercise is increasingly common but poorly reproducible. We highlighted that widespread inconsistency in methodologies may partly explain this variability [16]. To avoid misinforming the field, the quality of individual methodological approaches must be considered when integrating and discussing existing findings. The aim of the systematic review was to examine the current literature regarding the temporal response of cf-miRNA to an acute bout of exercise, and to interpret it in the light of the known limitations of the field using a robust multilevel meta-analysis.

## Methods

The systematic review was conducted and reported in accordance with the Preferred Reporting Items for Systematic Reviews and Meta-Analyses (PRISMA 2020, see supp file 1) [17]. The protocol was registered in the international prospective register of systematic reviews (PROSPERO: CRD42021256303). The Assessing the Methodological Quality of Systematic Reviews 2 (AMSTAR2) tool was used to appraise the systematic review (supp file 2).

### Search strategy

The electronic databases PubMed and SPORTDiscus were searched by one reviewer (KM) to identify all relevant articles and DH for the updated search. The detailed search criteria can be found in supp file 3. Databases were searched from inception to 17 May 2021 and updated on 12 August 2025. Additional articles were identified through hand searching the reference list of all included studies.

### Study inclusion and exclusion criteria

The Population, Intervention, Comparison, Outcomes and Study design (PICOS) framework was used to define the study eligibility criteria. Eligible participants were males and females who were: 18–45 yr; <30 kg/m^2^; and, free from known cardiac disease, metabolic disease or cancer. Studies including an acute exercise intervention of any duration and modality were eligible. Comparisons were made between relative miRNA expression levels pre and up to 48 hr post exercise bout. Studies including the primary or secondary outcome of cf-miRNA extracted from serum or plasma were eligible. Experimental studies were considered for analyses if they were original research articles, published in full in a peer reviewed journal and full text available in English. Limiting to English language has previously been shown to not meaningfully influence effect estimates [18].

### Study selection

Prior to study screening, all duplicate studies were removed. Duplicate studies were identified by conducting an automated title screen using excel and confirmed by manually screening the title, publication year and authors. Study selection was conducted independently by two reviewers (KM and DH, DH and SL [updated search]) and followed a two-phase screening strategy. In phase one, title and abstracts were screened in all studies identified during the electronic database search. Studies that did not meet the inclusion criteria were excluded. In phase two, full text screening was conducted to exclude all remaining articles that did not meet the inclusion criteria. Any disagreements in study eligibility between the two reviewers were discussed and resolved by consensus or by arbitration from a third reviewer (SL or CM).

### Data extraction

Data extracted from all eligible studies independently by at least two reviewers (KM, SB and DH) using an a-priori designed data extraction form. Data extracted included author information, participant characteristics, exercise type, sampling timepoints, and study outcomes. Additionally, methodological data regarding analysis of miRNA expression were extracted and included: method of miRNA expression analysis (i.e. single qPCR, miRNA array or via sequencing); miRNA species assessed; quality control for haemolysis, spike-in control for RNA extraction and reverse transcription and whether miRNA was normalised to spike-in or housekeeping miRNAs (supp file 4A for details). Any disagreements in data extraction between the two reviewers were discussed and resolved by consensus or by arbitration from a third reviewer (SL or CM). Where data were not available in the published manuscript, authors were contacted to obtain relevant information. Authors were emailed twice and given four weeks to respond. One article [19] was subsequently removed as the minimum data required for meta-analysis (mean and SD) could not be retrieved.

### Quality assessment

Quality of all included studies were assessed independently by two independent reviewers (KM and DH or DH and SL) using the risk of bias in non-randomised studies of interventions (ROBINS-I) tool [20]. Any disagreements between the two reviewers were discussed and resolved by consensus or by arbitration from a third reviewer (SL or CM). The ROBINS-I tool has seven domains: bias due to confounding, bias due to selection of participants, bias in classification of interventions, bias due to deviations from intended interventions, bias due to missing data, bias in measurement of outcomes and bias in selection of the reported results. Detailed information on the criteria used for grading are documented in supp file 5. Each domain was assessed as low, moderate, serious or critical risk of bias. The overall risk of bias for each study was determined to be the same level as the highest risk of bias allocated to an individual domain. Studies were not excluded from the meta-analysis based on risk of bias; however, risk of bias was taken into consideration during reporting of results. Results were visualised using the R package *robvis* [21].

### Certainty of evidence

The GRADEpro GDT software was used to assess certainty of evidence for primary studies [22]. This was based on risk of bias, inconsistency, indirectness, imprecision, and publication bias. The grades of evidence are characterised as follows: High certainty: we are very confident that the true effect lies close to that of the estimate of the effect. Moderate certainty: we are moderately confident in the effect estimate: the true effect is likely to be close to the estimate of the effect, but there is a possibility that it is substantially different. Low certainty: our confidence in the effect estimate is limited: the true effect may be substantially different from the estimate of the effect. Very low certainty: we have very little confidence in the effect estimate: the true effect is likely to be substantially different from the estimate of effect [23]. Detailed information on the criteria used for grading are documented in supp file 6.

### Data analyses

All data was analysed in R (v4.1.1). For each study, we computed the mean and standard deviation (SD) of the log fold-change (logFC) in miRNA expression, calculated via the delta-Ct method and expressed in arbitrary units, between the baseline and the relevant post-exercise timepoints. If minimum data relative mean or fold change or logFC and SD or SEM was not provided, authors were contacted for relevant information. Of the 34 eligible studies, 15 provided raw data, while the remaining 18 had data extracted from figures or tables. One study [19] was excluded due to insufficient data availability. To retain as much data as possible, time points were sorted into windows based on the following timepoints post-acute exercise: 0HP, 1– 2HP, 2–6HP, 6–24HP, > 24HP. Missing SD at baseline were imputed using k-nearest neighbours method implemented in the VIM package. We restricted analyses to miRNAs assessed in at least five studies per time window to minimise false-positive findings [24]. These included miR-1, miR-20a, miR-21, miR-126-3p, miR-126-5p, miR-133a/b, miR-146a, miR-206, miR-208b, miR-210, miR-221 miR-222, miR-378 and miR-499a. Timepoints were not evenly spaced and therefore we undertook sensitivity analysis to determine best-fitting correlation structure using a decay function that reduces correlation with increasing time lag. The variance-covariance matrix used in the meta-analysis was constructed using the best-fitting correlation parameters (ρ=0.6, decay rate=0.2). This matrix encodes the within-cohort covariance structure accounting for the decaying correlation between effect sizes measured at different timepoints. We then compared four different random-effects structures to identify the best model for capturing study- and cohort-level variability in miRNA responses to exercise using the *metafor* package [25].

1. Random Intercept Only: random = ∼ 1 | Study_ID.
2. Nested cohort model: random = ∼ 1 | Study_ID/cohort_id.
3. Random Slope Model (by Time) = ∼ Timepoint | Study_ID.
4. Nested Cohorts + Random Time Slopes =∼ 1 | Study_ID/cohort and ∼Timepoint | Study_ID

The best fit model was based on the lowest AIC (Supp File 4B). This was followed by cluster-robust estimate using the “sandwich” estimator to account for any misspecification of the model [26]. Statistical heterogeneity at each level of the meta-analysis was assessed via I^2^ which gives the proportion of total variability that is attributable to between-study heterogeneity rather than sampling error [27]. Tau^2^ (τ^2^) estimated the variability between studies, and 95% prediction intervals (PIs) were reported to reflect the expected range of true effects in future individual studies [28]. This provides a more conservative and informative interpretation of heterogeneity and the generalisability of the findings.

### Publication Bias and diagnostics

Publication bias was assessed visually via funnel plot asymmetry and tested using Egger’s regression was used to confirm the findings. Where there were less than 10 studies Egger’s regression test was not performed as there is a lack of power to detect reliable results [29]. Influential studies were identified through Cook’s distance (threshold >1) and outliers detected with studentised deleted residuals (>2) [30].

### Leave-One-Out and Sensitivity Analysis

Leave-One-Out (loo) and sensitivity analysis was conducted to assess the robustness of the meta-analysis findings. This analysis allowed us to assess how the exclusion of individual studies affected the overall effect estimate (intercept), heterogeneity (I^2^), and the statistical significance of timepoint-level moderators. A change of 5% in I^2^ and/or a change in significance at each timepoint was considered indicative of a meaningful impact on the model estimates.

### Exploratory/Meta-regression

To explore sources of heterogeneity, we examined eight moderators related to methodology, exercise modality, sample size, and participant sex. Given the large number of moderators relative to observations, we employed a machine-learning random forest approach (metaForest) to minimise the risk of overfitting the model to identify key moderators *[31]*. MetaForest models were built with 10,000 trees using random-effects weights clustered by cohort. Moderators with near-zero variance or low representation were excluded. Recursive variable selection via 100 bootstrap replications retained moderators selected in ≥50% of replications. Model convergence and predictive stability were assessed through 100 bootstrapped replications and R^2^ distributions.

### Narrative results

#### Literature search

The electronic database search retrieved 4318 articles, of which 1932 articles remained following removal of duplicates. Following title and abstract screening, 120 articles were identified as potentially eligible. Based on a full text assessment, 35 eligible articles were included in the meta-analysis (Figure 1).

**Figure 1.**
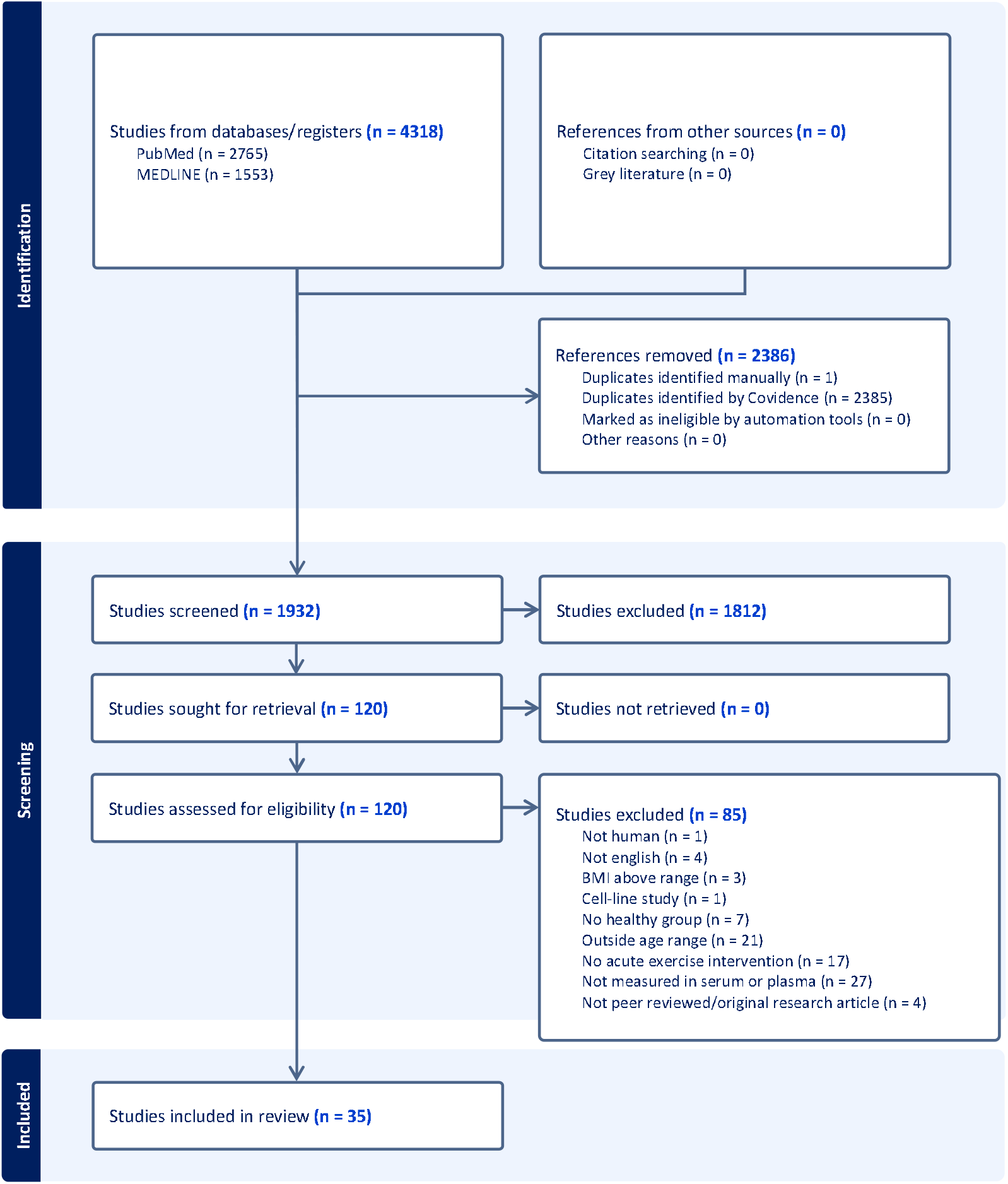
Preferred Reporting Items for Systematic Reviews and Meta-Analyses (PRISMA) guidelines flow chart for literature search and study selection.

#### Participant and study characteristics

Participant (N=736) and primary study (N=35) characteristics are described in Table 1.

**Table 1.** Study and participant characteristics of eligible studies.

| Combined Study Characteristics |  |  |  |
| --- | --- | --- | --- |
| Sample size range (N) |  | N = 4 – 120 |  |
| Age (Mean ± SD) |  | 24.2 ± 4.1 years |  |
| BMI (Mean ± SD) |  | 23.4 ± 2.1 kg/m<sup>2</sup> |  |
|  |  | Number of studies | Percentage |
| Male-only studies |  | 27/35 | 77% |
| Mixed studies |  | 8/35 | 23% |
| Female-only studies |  | 0/35 | 0% |
| Control of menstrual cycle |  | 1/8 | 12.5% |
| Training status | “Trained” | 16/35 | 46% |
|  | “Recreationally active” | 7/35 | 20% |
|  | “Untrained” | 8/35 | 23% |
|  | Not reported | 4/35 | 11% |
| Exercise modality | Endurance | 27/35 | 77% |
|  | Resistance | 8/35 | 23% |
| Timepoint | Immediately post-exercise | 32/35 | 91% |
|  | Between 1–12h | 22/35 | 63% |
|  | Between 13–48h | 12/35 | 34% |
| Tissue | Plasma | 22/35 | 63% |
|  | Serum | 13/35 | 37% |

#### MiRNA analysis characteristics

MiRNA analysis characteristics for primary studies (N=35) are described in Table 2.

**Table 2.** MiRNA analysis characteristics of primary studies.

| Quality control measures |  | Number of studies | Percentage |
| --- | --- | --- | --- |
| **miRNA expression analysis technique** | qPCR | 30/35 | 86% |
|  | Microarray | 4/35 | 11% |
|  | RNA sequencing | 1/35 | 3% |
| **Quality control for haemolysis\*** | Yes | 9/35 | 26% |
|  | No | 26/35 | 74% |
| **Quality control for RNA extraction (Spike-in)** | Yes | 23/35 | 66% |
|  | No | 11/35 | 31% |
|  | NA<sup>#</sup> | 1/35 | 3% |
| **Quality control for reverse transcription (spike-in)** | Yes | 7/35 | 20% |
|  | No | 27/35 | 77% |
|  | NA<sup>#</sup> | 1/35 | 3% |
| **Quality control for PCR (inter-plate calibrator)** | Yes | 4/35 | 11% |
|  | No | 30/35 | 86% |
|  | NA<sup>#</sup> | 1/35 | 3% |
| **Normalisation of miRNA expression<sup>^</sup>** | Yes | 35/35 | 100% |
|  | No | 0/35 | 0% |

\*Quality control for haemolysis included analysis of enriched blood miRNAs and/or spectrophotometry and/or visual inspection.
^ Normalisation of miRNA expression included use of a spike-in, endogenous miRNA(s) and/or global normalisation.

**Table 3:**
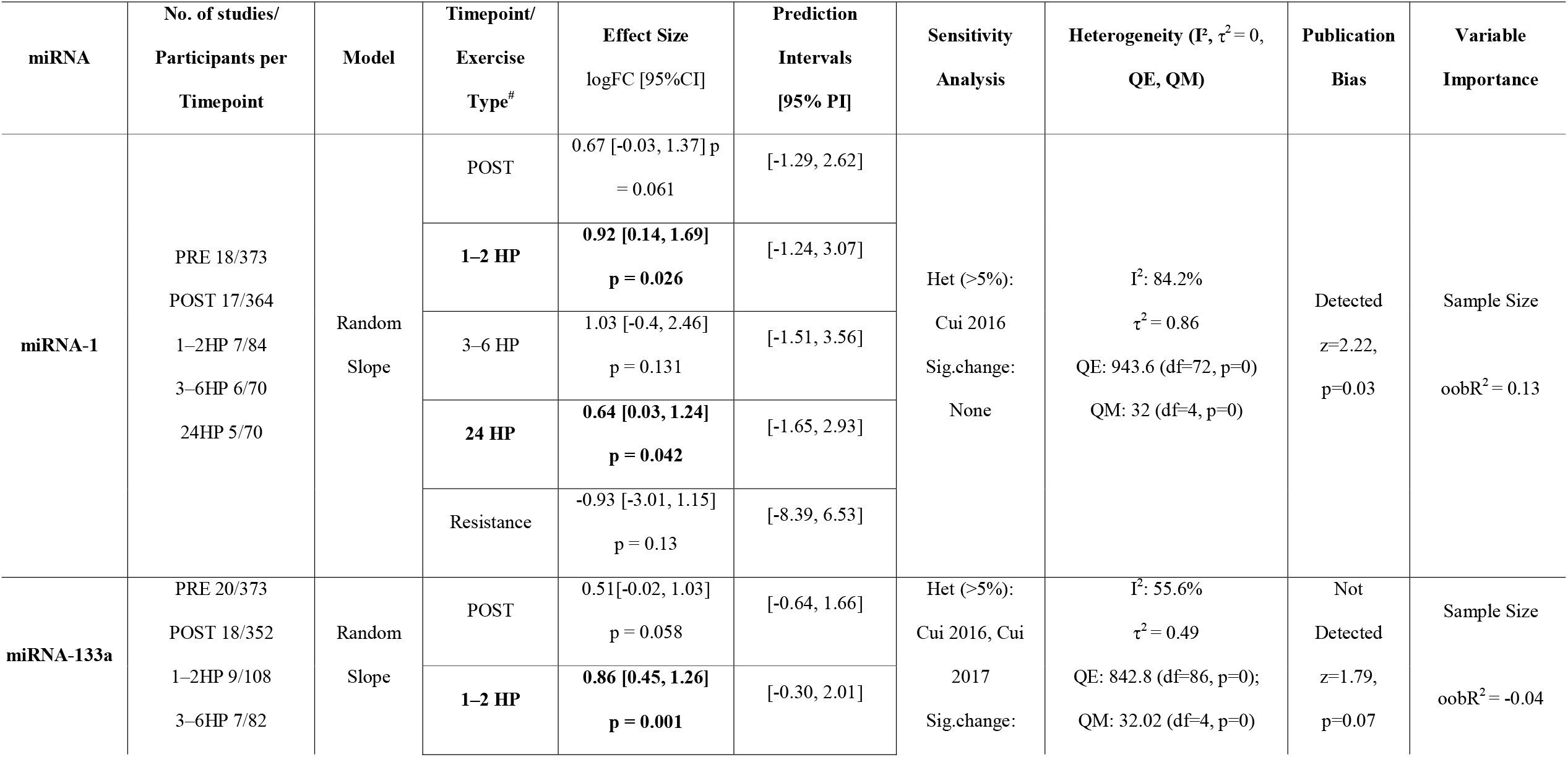
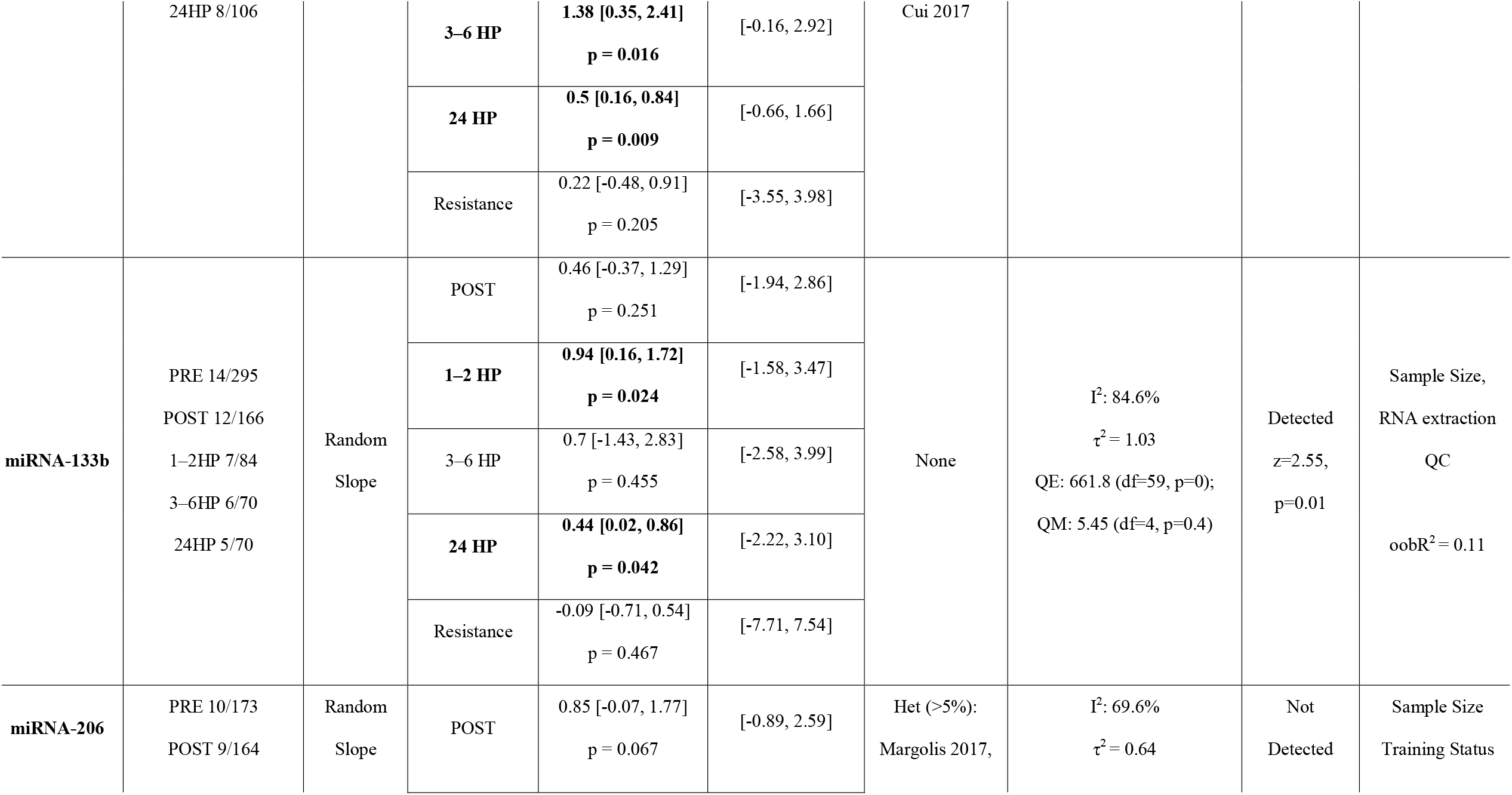
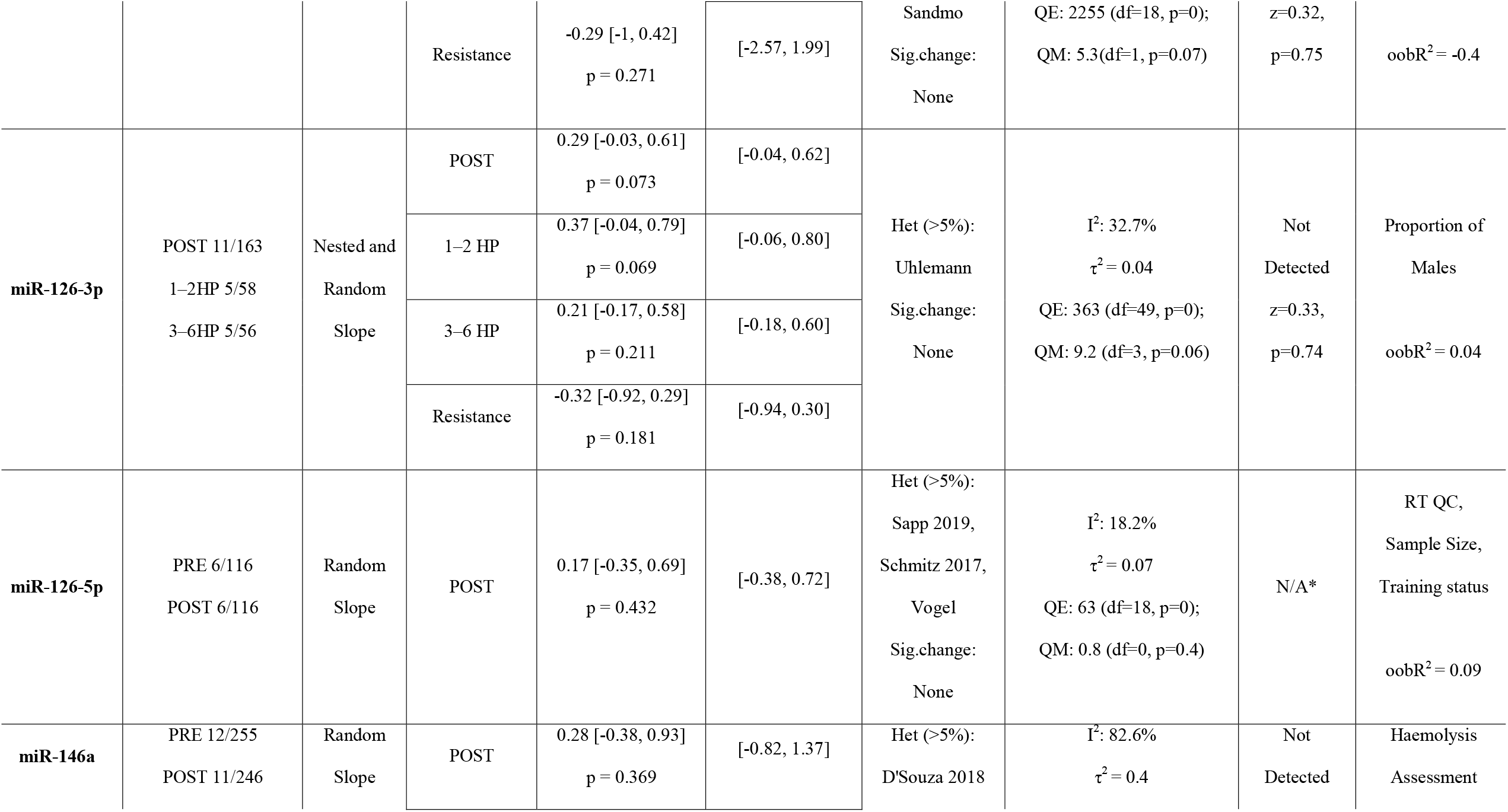
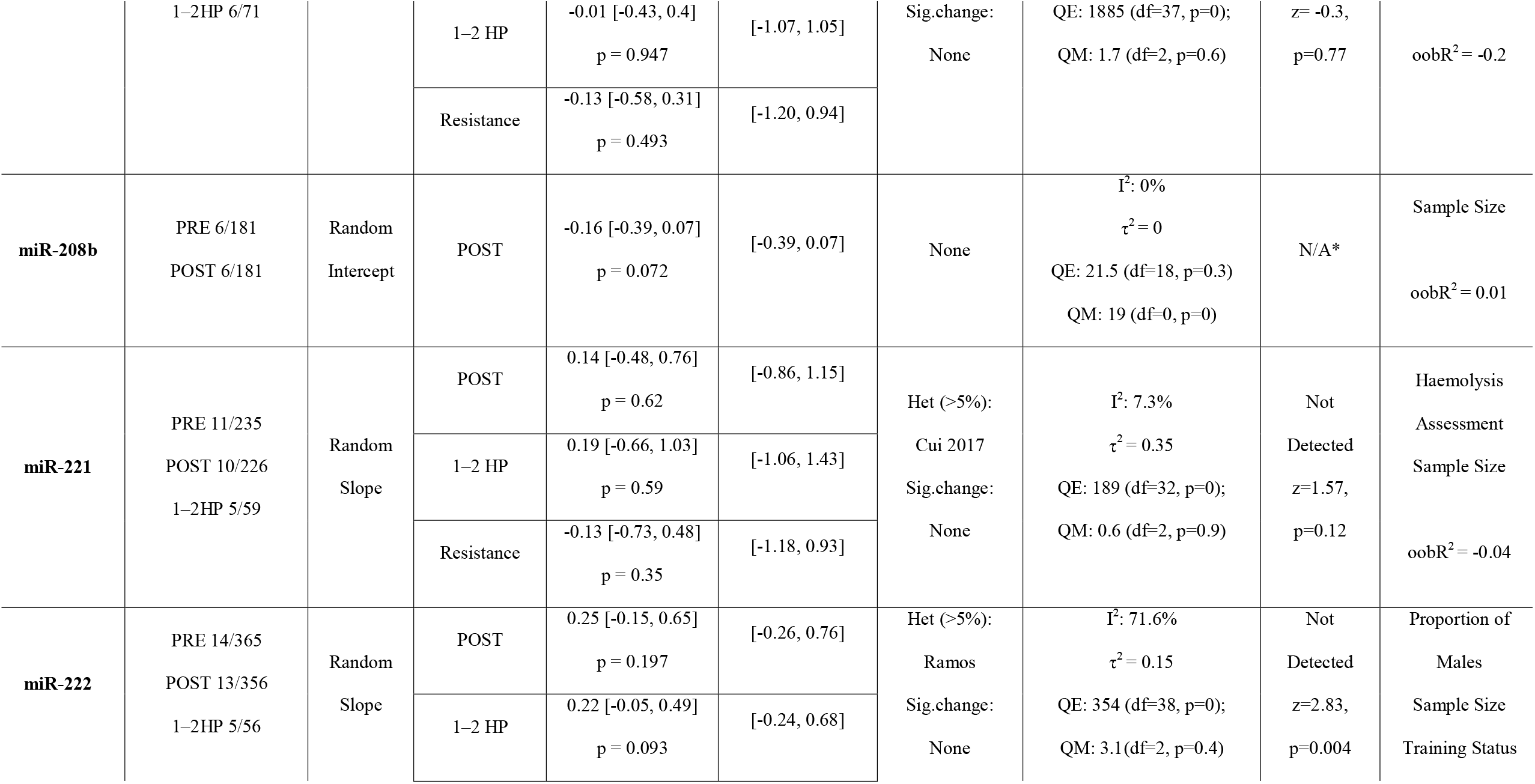
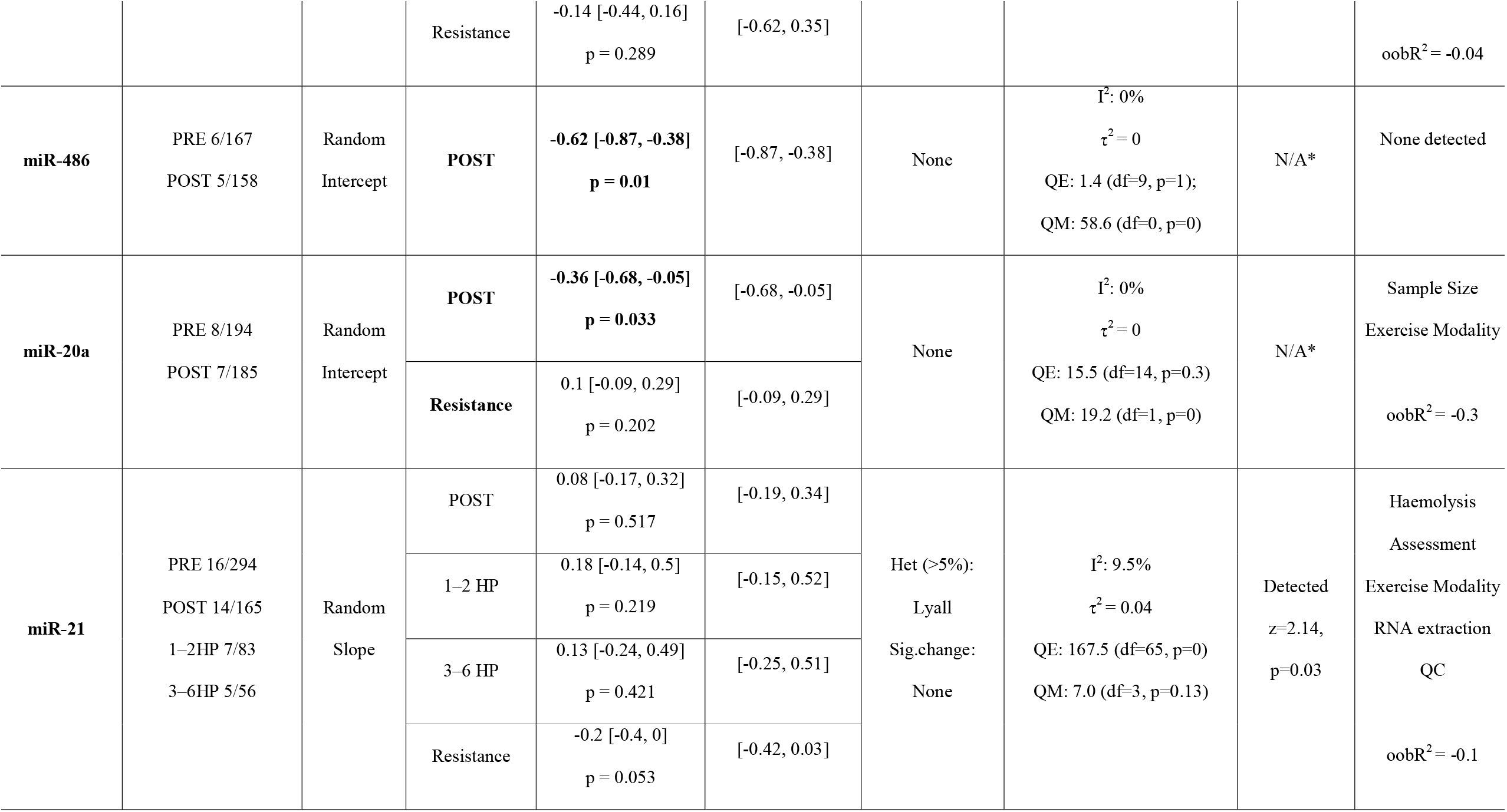
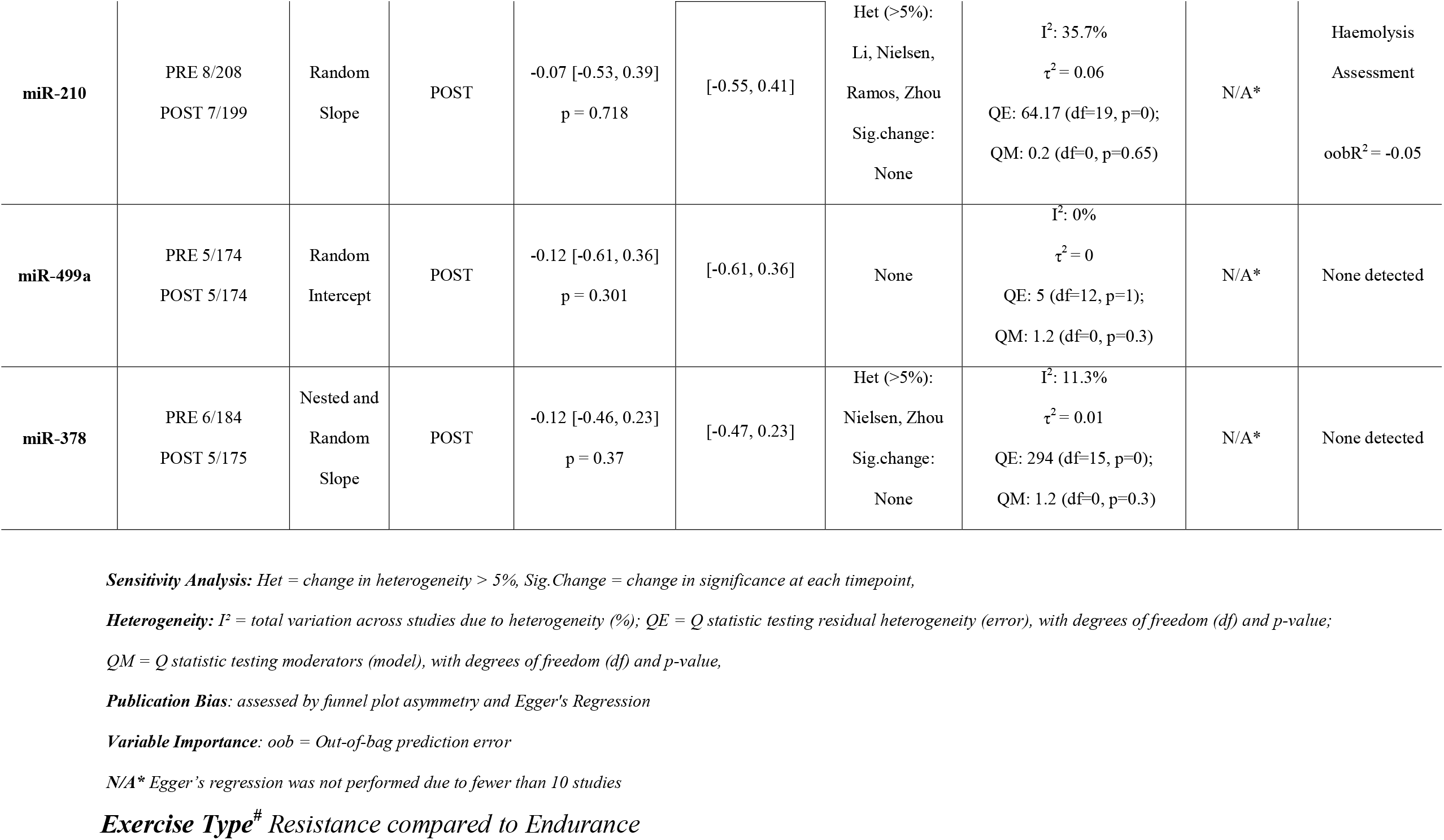
Summary of meta-analysis findings.

#### Study quality

Study quality was assessed using the ROBINS-1 tool for assessing risk of bias in non-randomised studies of interventions (supp figure 8). Quality of studies frequently decreased due to: failure to consider baseline confounding factors including age, body mass index (BMI), sex, menstrual cycle status and fitness in the study design; insufficient control for exercise intensity or duration; failure to appropriately control for nutritional intake or exercise 24 hr prior to the intervention; missing data without explanation and lack of reported N values; issues with reporting of miRNA statistical analysis, including absence of multiple testing correction and insufficient reporting of statistical analysis. Out of the 35 studies, two studies had an overall low risk of bias [32, 33], seven studies had an overall moderate risk of bias [34-40], 22 studies had an overall serious risk of bias [41-62] and 4 studies had an overall critical risk of bias [19, 63-65].

#### Certainty of evidence

Following GRADE assessment, there was a low certainty of evidence for all cf-miRNAs and our confidence in the effect estimate was limited. This was mainly due to a high risk of bias, and a high level of imprecision. These findings are summarised in supp file 6.

### Meta-analysis

#### Cf-miRNA-1-3p

Across 18 studies (N = 373), miR-1 levels significantly increased 1.9-fold at 1–2h and 1.6-fold 24h post-exercise but did not change immediately post or 3–6h post exercise (Figure 2A Table 1). There were no differences in miR-1 levels in response to endurance compared to resistance exercise. Sensitivity analyses identified the study by Cui et al. (2016) as a source of between-study variability but removing it did not alter the significance of the timepoint effects or the overall model estimates, indicating that the findings are robust. The analysis revealed substantial heterogeneity across studies (I^2^ = 84%; τ^2^ = 0.86), and wide prediction intervals reflect high uncertainty in the expected effect for future studies (Table 1). Moderator analyses identified sample size as the most notable variable explaining some of this heterogeneity, although it accounted for only a small proportion of the variance (OOB R^2^ = 0.013), indicating that most heterogeneity remains unexplained.

**Figure 2.**
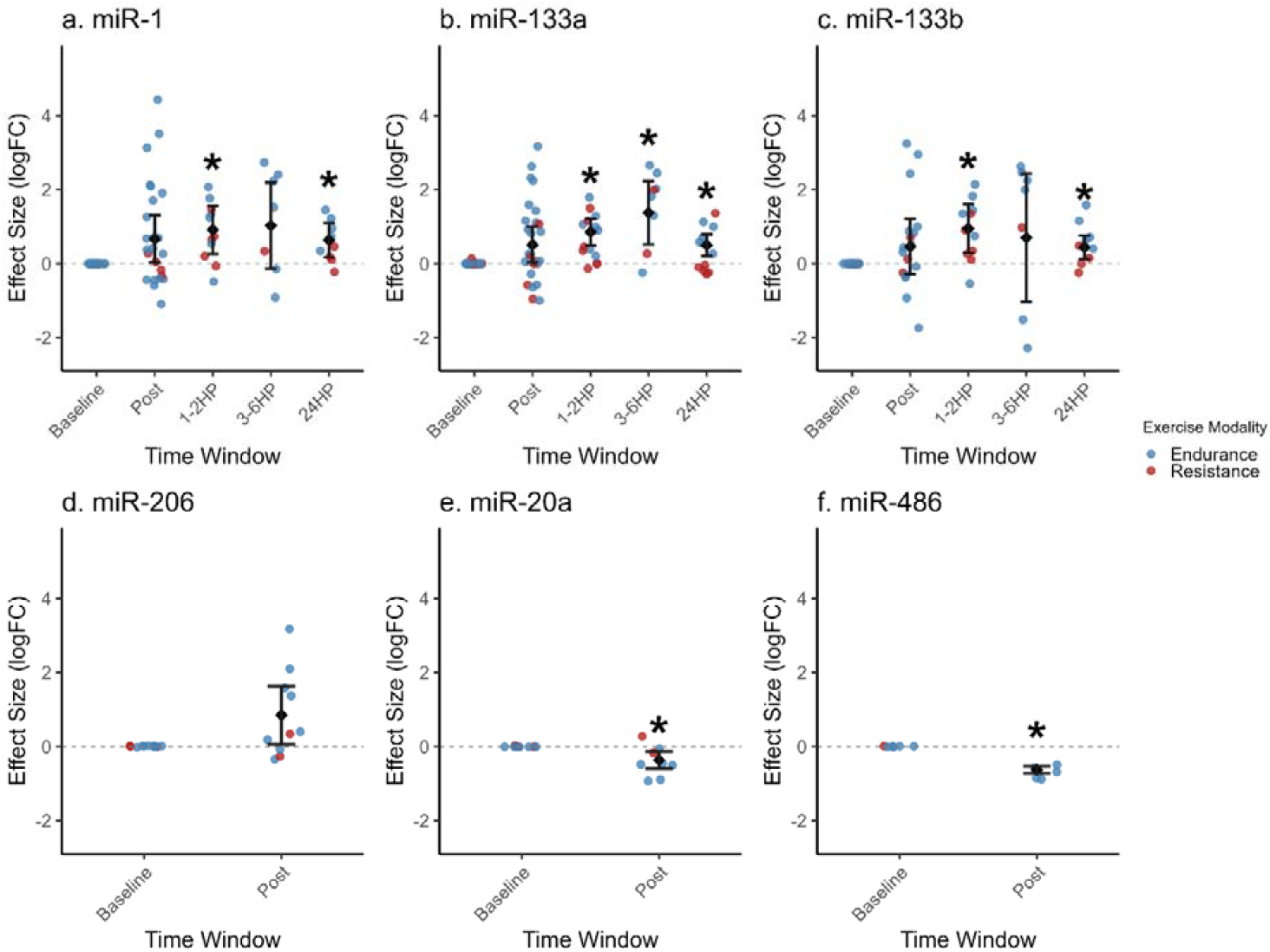
Individual study effects on miR-1 (a), miR-133a (b), miR-133b (c), miR-206 (d), miR-20a (e) and miR-486 (f) expression levels with meta-analysis estimates (logFC) and 95% confidence intervals. *Significant difference compared to baseline p<0.05.

#### Cf-miRNA-133a

Across 20 studies (N = 373), miR-133a levels significantly increased 1.8-fold at 1–2h, 2.6-fold at 3–6h, and 1.4-fold at 24h post-exercise, with no influence of exercise modality (Figure 2B Table 1). Heterogeneity across studies was moderate (I^2^ = 48%, τ^2^ = 0.49), with two studies (Cui 2016 and Cui 2017) significantly contributing to this heterogeneity. Leave-one-out sensitivity analyses identified Cui (2017) as influential, with the exclusion of this study affecting the significance of the post-exercise timepoint effects. Additionally, wide prediction intervals indicate that future studies may observe variable miR-133a responses (Table 1). Overall, miR-133a demonstrates more consistent post-exercise increases compared to miR-1. Moderator analyses again indicated that sample size was the only notable variable that influenced between-study heterogeneity. However, the overall model fit was poor (OOB R^2^ = –0.04), suggesting that sample size only explained a small amount of the variability observed.

#### Cf-miRNA-133b

Across 14 studies (N = 295), miR-133b increased 1.9-fold in the 1–2h post-exercise and 1.4-fold at 24h post-exercise but did not change immediately post or 3–6h post-exercise (Figure 2C Table 1). Heterogeneity was substantial (I^2^ = 85%, τ^2^ = 1.03), though sensitivity analyses did not identify any single study as disproportionately influential. Wide prediction intervals indicate once again that future studies may observe variable miR-133b responses (Table 1). Moderator analyses suggested that sample size and the inclusion of a spike-in control to normalise for differences in miRNA input during RNA extraction (Extraction_QC) were the most influential variables associated with between-study variability.

#### Cf-miRNA-206

Findings from the 10 eligible studies having studied miR-206 (N = 173) are summarised in Figure 2D Table 1. There were no differences in miR-206 levels immediately following a bout of acute exercise, the only time point investigated. Exercise modality did not influence miR-206 levels. Between-study heterogeneity was moderate (I^2^ = 70%, τ^2^ = 0.64) and wide prediction intervals indicate that future studies may observe variable miR-206 responses (Table 1). Leave-one-out sensitivity analysis showed that two studies (Sandmo et al., Margolis (2017)) meaningfully influenced heterogeneity (Δ I^2^ > 5%). However, the exclusion of any single study did not affect the statistical significance of the intercept or timepoint moderators. Moderator analysis revealed that sample size and training status were the most important variables; however, poor overall model fit (OOB R^2^ = – 0.4) indicates these moderators only explained a small amount of the variability observed.

#### Cf-miRNA-20a and Cf-miRNA-486

Across 8 studies (N = 194) and 6 studies (N = 167), miR-20a and miR-486 decreased 0.7-fold and 0.65-fold immediately post-exercise, respectively (Figure 2E and 2F, Table 1). Exercise modality did not influence miR-20a levels. Between-study heterogeneity was null (I^2^ = 0%, τ^2^ = 0) for both miR-20a and miR-486 and a fixed effect model provided a slightly better fit than the random effect models, reflecting limited between study-heterogeneity. This consistency across studies supports the reliability of the pooled effect estimate. Further, the prediction interval was relatively narrow, indicating consistent decreases in miR-20a and miR-486 levels across studies post-exercise and suggesting future studies are likely to observe similar results with less variability (Table 1). Moderator analysis of miR-20a revealed that sample size and exercise modality were the most important variables; however, poor overall model fit (OOB R^2^ = –0.3) indicates these moderators explained little of the between-study variability observed.

#### Cell-free miR-126-3p, miR-126-5p, miR-146a, miR-21, miR-210, miR-221, miR-222, miR-378, miR-499a, miR-208b

None of the following miRNAs (miR-126-3p, miR-126-5p, miR-146a, miR-21, miR-208b, miR-210, miR-221, miR-222, miR-378, miR-499a) investigated across 5–14 studies per miRNA (sample size ranged between 98– 361 participants; see Table 2 for individual study details) showed significant changes immediately or at any other timepoints post-exercise. Between-study heterogeneity ranged from low (I^2^ ∼7%, τ^2^ = 0.001) to substantial (I^2^ up to 82%, τ^2^ up to 0.4). Correspondingly, prediction intervals ranged from relatively narrow to very wide, reflecting differences in the precision and consistency of the effect estimates and suggesting variable responses in future studies (Table 1). Sensitivity analyses identified some studies as influential; however, only in the miR-126-3p meta-analysis did the exclusion of the study by Margolis (2022) lead to a statistically significant change at the post-exercise timepoint. Moderator analyses highlighted variables such as haemolysis assessment, sex, sample size, training status, and RNA extraction quality as the most commonly important moderators. However, model fits were generally poor (OOB R^2^ typically ≤ 0), indicating that these moderators explained only a small fraction of the variability.

## Discussion

The primary aim of this systematic review and meta-analysis was to evaluate the influence of an acute exercise bout on cf-miRNA levels in apparently healthy individuals. Sixteen individual cf-miRNAs across 35 primary studies (participants n= 736) met our inclusion criteria. Our main findings were that cf-miR-133a levels increased 1–2 hr, 3–6 hr and 24 hr post exercise, while cf-miR-1 and cf-miR-133b levels increased 1–2 hr and 24hr post exercise, but not 3–6hr post exercise. Cf-miR-20a and cf-miR-486 were downregulated immediately post exercise. Collectively, our findings reveal temporal cf-miRNA responses to an acute exercise bout with little influence of exercise modality.

### Muscle-enriched cell free miRNAs levels are altered with acute exercise

Exercise is one of the most potent physiological stimuli. Muscles contract and relax repeatedly, leading to substantial changes in multiple physiological systems. A recent meta-analysis showed that out of the ∼20,000 human protein coding genes [66], almost 3,000 genes change their expression in response to acute exercise [67]. The human genome also encodes non-protein coding genes, including over 2,600 mature miRNAs (miRBase v. 22), with at least 10–15% exported out of the cell and found at detectable levels in the circulation [68]. Cf-miRNAs may be involved in cell-cell communication, influencing gene expression in other cells rather than the one that produced them [14, 15]. Over 60% of human protein coding genes may be regulated by miRNAs [69], meaning that every signalling pathway likely contains some elements that are under miRNA regulation. Most miRNAs can bind to multiple mRNAs [70], and each mRNA typically has binding sites for several distinct miRNAs; although, in both cases, the distribution is not uniform [71]. This network of interactions allows miRNAs to function in groups to fine tune control of gene expression with a high degree of precision. The specific miRNAs reported here were chosen due to their investigation in at least five primary studies. Aside from cf-miR-20a, all miRNAs that were altered with exercise were those originally classed as “myo-miRs” [72], suggesting that they may work together. Their presence in the circulation suggests that they are exported from exercising muscle and may be signalling to other tissues to prepare for physiological changes [73].

Canonical myomiRs miR-1 and miR-133a/b are widely conserved miRNA showing high levels of expression primarily in muscle tissue [74]. The genes that produce miR-133a and miR-1 are clustered on the same chromosomal loci and are transcribed together [75]. MiR-1 is a key regulator of skeletal muscle response to exercise regulated by mTOR, possibly via modulation of the IGF-1/PI3K/AKT and IGF-1/IGF-1R pathways [76-78]. MiR-133a stimulates skeletal muscle growth by reducing myoblast differentiation, with downregulation of tropomyosin-4, an actin binding component of the cytoskeleton [79] or predicted binding sites in the TCF7, MSI1, and PAX5 genes of the Wnt signalling pathway [80] as possible mechanisms. MiR-133b is identical to miR-133a except for a single base at the 3’ end and is similarly enriched in muscle tissue [75]. Cf-miR-133b is reduced in sarcopenic individuals [81], and like miR-133a, targets the Wnt signalling pathway [82]. Several studies have shown that the secretion of these miRNAs into circulation is a tightly regulated and selective process [83, 84]. For example, a recent study showed that miR-1 is secreted from the muscle following an acute bout of resistance exercise and transferred to adipose tissue via extracellular vesicles, where it was shown putatively to reduce the expression of *CAV2* and *TRIM6* which are linked to lipolysis [85]. Therefore, the elevation of cf-miR-1, -133a, and -133b in circulation post-exercise may result from the muscle tissue undergoing remodelling, with cf-miRNAs serving as signalling molecules that potentially coordinate systemic responses within other tissues [73].

Although miR-206 is also a muscle-enriched myo-miR transcribed from the same gene as miR-133b, it did not show significant changes in circulation following exercise in our study, suggesting a distinct regulatory role or release mechanism compared to miR-1 and miR-133a/b. MiR-206 is primarily involved in stem cell and myoblast differentiation within muscle tissue [86, 87] and therefore may not show an immediate increase post-exercise [88].

MiR-486, another muscle-enriched miRNA, is regulated by myocardin and myocardin-related transcription factors (MRTFs), serum response factor (SRF) and MyoD [89]. It plays a key role in muscle differentiation, proliferation, and regeneration by regulating tensin homolog (PTEN) and FOXO1a and enhancing the expression PI3K/AKT signalling pathway [89, 90]. Unlike cf-miR-1, miR-133a, and miR-133b, cf-miR-486 decreased following exercise, which may reflect increased uptake/production into muscle tissue where it may regulate repair or regeneration [91]. Overall, these opposing directional change reflect distinct roles, as miR-486 might be retained in the tissue to mediate intracellular pathways, while miR-1, miR-133a, and miR-133b may be actively secreted to act as intercellular messengers modulating pathways in other tissues[83].

Aspects of the methodological design (e.g. normalisation method), sample size and proportion of males are common pitfalls in cf-miRNA studies [16, 92, 93]. An exploratory machine-learning-based approach identified sample size as the highest ranked moderator that may be influencing effect size in the analysis of miRNA-1, miRNA-133a, miRNA-133b, miR-206, miR-126-5p, miR-208b, miR-221, miR-222 and miR-20a. This is not overly surprising as small sample size in primary studies is a commonly cited limitation of meta-analyses that leads to increased risk of type I errors (false negatives) and type II errors (false positives) in the overall effect size [94]. Along the same lines, implementation of best practice recommendations for miRNA analysis [16, 38], including stringent and rigorous methodological checks, was commonly missing and may also explain some of the variability in effect sizes. Specifically, the assessment of haemolysis (missing in 74% of studies) and inclusion of a spike-in control to normalise for differences in miRNA input during RNA extraction (missing in 31% of studies) were identified as influential moderators. Failure to control for haemolysis can cause discrepancies between studies [16] as erythrocytes can contain miRNAs that may confound results if not properly eliminated [16, 95]. Due to the difficultly to accurately determine the concentrations of cf-miRNAs, many studies elect to use equal amounts of total RNA volume as a surrogate for equal amounts of RNA input for RNA extraction [16]. However, this method is only valid if an appropriate spike-in control is used to account for differences in cf-miRNA input. Altogether, these differences in the use of quality controls can dramatically increase between-study variability. As previously discussed by our group [16], these methodological checks should become the ‘norm’ in cf-miRNA studies as they increase reproducibility and will lead to a more robust understanding of the cf-miRNA response following exercise as well as in multiple pathophysiological conditions.

Given the known differences in physiological demand that result from different modes of exercise [96], we hypothesised that individual miRNAs may respond differently to different acute exercise protocols. However, we did not detect differences in the overall expression levels of the included miRNAs between exercise modality groups. It should be noted that, due to the limited number of primary studies, we were unable to run an interaction analysis to delineate the influence (if any) of exercise modality on the levels of these miRNAs at each individual timepoint. It will be important to investigate these differences between exercise modalities in future studies to understand if and how different modes of exercise can influence cf-miRNA levels, and therefore the specific pathways that are altered.

Consistent with previous reports in the fields of physiology [97] and exercise physiology [98], only 23% of the 35 primary studies included female participants, with none being female-only. Moreover, menstrual status was controlled in only one of these studies [99]. Historically, sex-specific differences in exercise response and other fields of muscle physiology and medicine have been overlooked due to financial costs and feasibility [100]. However, this lack of female-only and mixed-sex studies leads to insufficient information to differentiate between the divergent exercise response of males and females [101-104]. Our group’s research revealed profound differences in the skeletal muscle miRNAome of males and females at baseline and 3 hours post an acute exercise bout [103]. Furthermore, there is literature examining sex-specific differences in miRNA expression in other tissues [105-107] and plasma [108], where androgenic and ovarian hormones may modulate the expression of specific miRNA species [108-110]. Future studies should include sufficient numbers of participants of both sexes to assess sex as a factor in their analyses. Sex should be retained as a fixed factor or covariate when it meaningfully contributes to explaining variability or reduces the residual variance or slightly improves model fit (even if the effect is not statistically significant). This approach balances simplicity with appropriate adjustment for biological differences, avoiding unnecessary complexity while ensuring the robustness of findings.

## Strengths and limitations

This systematic review included 35 primary studies; however, few were considered low risk of bias. Bias due to deviations from intended interventions and co-interventions were judged moderate, serious or critical in 31 of the 35 primary studies. This was primarily attributed to insufficient control of the exercise intervention, including intensity and duration, and nutritional intake. Another, common limitation was the lack of appropriate reporting or control of baseline confounding variables, which may however be less consequential in randomised cross-over or repeated-measures designs where each individual acts as their own control. Many studies did not report or account for key factors, such as fitness level, BMI, sex, and age, all of which are well-established moderators of miRNA expression [42, 103, 111]. Future studies within the field could implement rigorous methodology to minimise risk of bias, increase robustness and reproducibility, and generally facilitate comparability within the literature body. Improving quality, rigour and reproducibility [112] in physiological sciences has recently become a focus of many [113, 114]. As we hope to increase reproducibility and transparency in the cf-miRNA field [16], the full R code and the data used in the analysis is available on GitHub (https://github.com/DaniHiam/circulating-miRNAs-and-exercise-).

In conclusion, we reported the effect of an acute bout of exercise on common cf-miRNAs in humans. We conducted a comprehensive longitudinal meta-analysis across 16 cf-miRNAs followed by machine-learning approach to identify key methodological factors influencing their regulation. Our findings demonstrate that muscle-enriched cf-miRNAs are dynamically regulated post-exercise, displaying distinct temporal expression patterns, and provide critical insights to guide future research on cf-miRNA dynamics in exercise contexts. However, given the limited sample sizes and variability in methodological quality across the reviewed studies, the overall evidence remains preliminary. Future work should prioritise improving study design and standardising methodologies to enhance the robustness and reproducibility of findings.

## Data Availability

The full R code and the data used in the analysis is available on GitHub (https://github.com/DaniHiam/circulating-miRNAs-and-exercise-).

## Notes

**Funding:** SL is supported by Australian Research Council (ARC) Future Fellowship

### Competing Interest Statement

Associate Professor Patrick J Owen is an Editorial Board Member at Sports Medicine but was not involved in the selection of peer reviewers for this manuscript or any of the subsequent editorial decisions.

### Funding Statement

This study did not receive any funding

### Author Declarations

The electronic databases PubMed and SPORTDiscus were searched to identify all relevant articles

### Summary of Updates

The statistical methodology has been revised, the literature search updated, and the discussion expanded.

