## Supplementary material for "Response of cell-free miRNAs to acute exercise: A systematic Review and Meta-Analysis": Supp File 3 - Search Terms.pdf

**Search strategy**

| Keywords | Search term | Limits |
|---|---|---|
| MicroRNA | microRNA* OR "micro RNA*" OR miRNA* OR miR | • English language<br>• Search terms in title/abstract (PubMed)<br>• Journal articles<br>• Humans |
| Exercise | exercis* OR "exercise therapy" OR "physical activity" OR training OR endurance OR aerobic, resistance OR strength OR weight* OR isometric | |
| Circulating | Circulat* OR plasma OR serum OR "cell-free" OR "cell free" OR exosom* OR vesicular OR "extracellular vesicle" OR extracellular | |
