## Supplementary material for "Response of cell-free miRNAs to acute exercise: A systematic Review and Meta-Analysis": Supp File 5 - ROBINS-1 criteria.pdf

**ROBINS-I-v2 Tool**

Domain 1- Confounding:

- Baseline confounding: Can we say they have a similar baseline based on following characteristics:
  - Age,
  - BMI,
  - Sex,
  - Menstrual cycle (if female),
  - Fitness status.
- Time varying confounding, based on the following:
  - Fasting duration of > 8 hr.
  - Diet control
  - Exercise controlled 24 hr prior to testing

Domain 2- Classification of intervention:

- Did assignment to a group rely on measurements post-enrollment?
- Was classification based on pre-intervention information?
- Was classification influenced by knowing the outcome or risk?
- As a randomised crossover with balanced order and clear protocol, misclassification of assigned intervention is highly unlikely.
- Study provides clear details of the following exercise intervention parameters:
  - Type
  - Dose
  - Frequency
  - Intensity and/or duration

Domain 3- Selection of participants:

- Was selection into study based on participant characteristics after start of intervention.
- Given the nature of included studies intervention's, most studies will be LOW for domain.
- Excluding participants based on serum or miRNA QC are irrelevant in this domain, as per guidance doc- "selection bias occurs when selection of participants is related to both intervention and outcome".

Domain 4- Deviations from intended interventions:

- Was there sufficient control for implementation of the intended treatment protocol? Consider whether there was sufficient control of the following:

  - Exercise intensity (Rate of Perceived Exertion (RPE), Heart Rate, or intensity controlled between participants).
  - Exercise duration.
  - Time of day participants

- Co-interventions that apply to pre and post intervention timepoint. Consider the following:
  - Fasting duration of > 8 hr or controlled diet night before or morning of intervention
  - Exercise controlled 24 hr prior to testing.
- Are N values provided to confirm the intervention was successful in all participants?

<u>Domain 5- Missing data:</u>

- Consider the following:
  - Was there any missing data?
  - Were N values provided in tables/graphs?
  - Were reasons provided for missing data?
  - How was missing data dealt with?

<u>Domain 6- Measurement of outcomes:</u>

- Where outcome measures influenced by knowledge of intervention (i.e timepoint?)
- Was the blood sampling timepoint and time of collection similar between all participants?
- Due to the objective nature of assessment of miRNA expression, most studies will score low for this domain.

<u>Domain 7- Selected of reported result:</u>

- Was there selective reporting of results (did they run the stats in multiple way to get the "best" outcome)? Consider the following:
  - Have they stated the same technique in the methods as is reported in the results? (e.g. ddCt).
  - How were housekeeping genes selected? A priori? Using statistical test (e.g. normfinder)
  - Were corrections made for multiple analysis? (e.g. FDR).
