## Supplementary material for "Response of cell-free miRNAs to acute exercise: A systematic Review and Meta-Analysis": Supp File 6 - GRADE Assessment.pdf

| Outcomes | № of studies | Certainty of the evidence (GRADE) |
| --- | --- | --- |
| miRNA-1 | (18 non-randomised studies) | ⊕◯◯◯ Very low[a,b,c,d,e,f] |
| miRNA-133a | (20 non-randomised studies) | ⊕◯◯◯ Very low[a,b,c,d] |
| miRNA-133b | (14 non-randomised studies) | ⊕◯◯◯ Very low[a,b,c,d,e,f] |
| miRNA-206 | (10 non-randomised studies) | ⊕◯◯◯ Very low[a,b,c,d,e] |
| miRNA-146a | (12 non-randomised studies) | ⊕◯◯◯ Very low[a,b,c,d,e] |
| miRNA-20a | (8 non-randomised studies) | ⊕◯◯◯ Very low[a,d] |
| miRNA-21 | (16 non-randomised studies) | ⊕◯◯◯ Very low[a,f] |
| miRNA-126-3p | (12 non-randomised studies) | ⊕◯◯◯ Very low[a,d,e] |
| miRNA-126-5p | (6 non-randomised studies) | ⊕◯◯◯ Very low[a,d,e,f] |
| miRNA-208b | (6 non-randomised studies) | ⊕◯◯◯ Very low[a,d,e] |
| miRNA-210 | (8 non-randomised studies) | ⊕◯◯◯ Very low[a,e] |
| miRNA-221 | (11 non-randomised studies) | ⊕◯◯◯ Very low[a,d,e] |
| miRNA-222 | (14 non-randomised studies) | ⊕◯◯◯ Very low[a,b,c,d,e] |
| miRNA-378 | (6 non-randomised studies) | ⊕◯◯◯ Very low[a,c,d,e] |
| miRNA-486 | (6 non-randomised studies) | ⊕◯◯◯ Very low[a,d] |

| Outcomes | № of studies | Certainty of the evidence (GRADE) |
|---|---|---|
| miRNA-499a | (5 non-randomised studies) | ⨁◯◯◯ Very low[a,d,e,f] |

**Explanations**

a. Based on ROBINS-I tool (Multiple studies at serious/critical risk of bias)

b. Substantial unexplained heterogeneity ($I^2 > 50\%$, $p < 0.05$) and could not be explained by meta regression or sensitivity analyses.

c. Sensitivity Analysis (change of 5% points in $I^2$ and/or a change in significance at each timepoint)

d. No downgrade for indirectness as all included studies matched the pre-defined PICO criteria

e. Imprecision due to wide 95% confidence intervals, reflecting uncertainty in the magnitude and direction of the effect.

f. Asymmetry detected in the funnel plot and a significant eggers regression test.

**GRADE Working Group grades of evidence**

**High certainty:** we are very confident that the true effect lies close to that of the estimate of the effect.

**Moderate certainty:** we are moderately confident in the effect estimate: the true effect is likely to be close to the estimate of the effect, but there is a possibility that it is substantially different.

**Low certainty:** our confidence in the effect estimate is limited: the true effect may be substantially different from the estimate of the effect.

**Very low certainty:** we have very little confidence in the effect estimate: the true effect is likely to be substantially different from the estimate of effect.
