## Supplementary figures and images for "Response of cell-free miRNAs to acute exercise: A systematic Review and Meta-Analysis"

### Supp File 8- ROBINS-I figure.jpeg

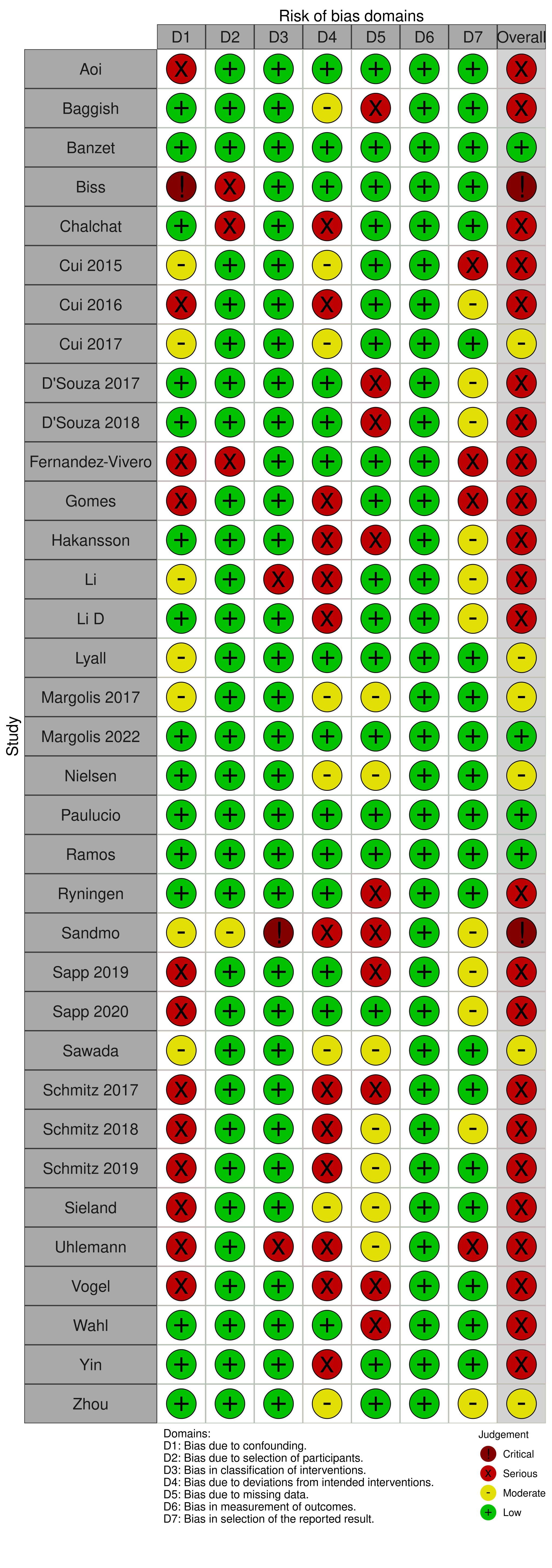
